# Therapeutic effects of PTCD and ERCP in patients with obstructive severe acute biliary pancreatitis

**DOI:** 10.1101/2022.04.29.22274457

**Authors:** Xue Ling Zhang, Jia Huan Sun, Yue Wu, Min Xie, Cong Cong Li, Dong Lv, Wei Yu, Pei Lin Cui

## Abstract

**Objective:** We evaluated the therapeutic effects of PTCD and ERCP in patients with obstructive severe acute biliary pancreatitis (SABP).

**Methods:** A total of 62 patients with obstructive SABP were enrolled in this study from July 2013 to July 2019 and divided into three groups: PTCD group (n=22), ERCP group (n=24) and conservative group (n=16). Based on treatment time, PTCD and ERCP groups were further separated into early (⩽ 72 h) and delayed (>72 h) groups. Laboratory indices, hospitalization days, recovery of liver functions and remission of abdominal pain as well as complications were evaluated to establish the efficacy and suitable time.

**Results:** The average hospitalization days, time for abdominal pain relief and laboratory indices (leukocyte, blood amylase, ALT and TBiL) recovery were shorter (*p*<0.05) in PTCD and ERCP groups. The average hospitalization days for the ERCP group (16.71±3.99) were shorter, compared to the PTCD group (19.64±4.27) (*p*<0.05). Complications were few in ERCP (33.33%) and PTCD (27.27%) groups. The average length of stay (13.88±3.27), recovery time of leukocyte (6.31±0.92) and TBiL (9.13±1.98) in the early ERCP group were shorter than in both delayed ERCP and early PTCD groups (*p*<0.05). The average length of stay (18.63±4.06) and ALT recovery time (12.25±2.59) in delayed ERCP group were shorter than in delayed PTCD group (*p*<0.05).

**Conclusions:** Both ERCP and PTCD are effective for relieving biliary obstruction during SABP, and early ERCP or PTCD within 72 h for obstructive SABP are more beneficial.

## 1. Introduction

Acute pancreatitis (AP) is a common clinical emergency and a heterogeneous condition with the potential for significant morbidity and mortality^[1]^. In the past few years, incidences of first-time acute pancreatitis and disease-related complications have been increasing^[2]^. Acute biliary pancreatitis (ABP) accounts for 50%-70% of acute pancreatitis incidences^[3]^. The etiology of ABP is complex and multifactorial. Gallstones are the main cause of acute pancreatitis^[4,5]^. Severe acute pancreatitis is characterized by persistent single or multiple organ failure, which is associated with mortality rates between 20% and 40%^[6-9]^. About 20% of acute pancreatitis patients exhibit severe acute pancreatitis episodes. Determinant-based classification^[10]^ (DBC) and revised Atlanta classification^[7]^ (RAC) support severe acute pancreatitis is characterized by local or systemic complications and persistent organ failure (POF^[11]^>48 h) (cardiovascular, respiratory, and renal system).

Even though early management of different degrees and reasons of pancreatitis is majorly focused on fluid resuscitation, removal of biliary obstruction is advocated for obstructive severe acute biliary pancreatitis (SABP)^[12]^. Biliary obstruction can be removed via endoscopic retrograde cholangiopancreatography (ERCP), percutaneous transhepatic cholangial drainage (PTCD) and surgical treatment. ERCP is the primary option for ABP patients with acute cholangitis^[13]^; however, the 2013 IAP/APA guidelines^[14]^ suggest that there is no evidence for optimal timing of ERCP treatment for biliary pancreatitis patients without cholangitis. With regards to treatment timing of ERCP, the 2015 Italian consensus guidelines^[11,15]^ on severe acute pancreatitis recommend that when biliary obstruction is confirmed, ERCP should be performed within 72 h of admission. However, the 2019 ASGE guidelines^[16]^ on the role of endoscopy in evaluation and management of choledocholithiasis suggest the emergency (within 48 h) ERCP is recommended for biliary pancreatitis patients with biliary obstruction or bile duct stones. The guidelines and meta-analyses data show that a suitable time for endoscopy treatment of severe acute biliary pancreatitis is incoherent.

PTCD is widely accepted as an alternative to operative decompression in patients with cholangitis or cholecystitis, particularly elderly patients^[17-19]^ and SABP patients who cannot tolerate endoscopy^[20]^ or for whom endoscopy is unsafe^[21]^. A retrospective study^[22]^ involving 64 patients with obstructive SABP revealed that laboratory indicators and APACHE-II scores were decreased after early PTCD. However, there is insufficient evidence to support the effects of ERCP and PTCD in improving the clinical efficacies, such as laboratory indices, hospitalization stays, recovery of liver functions, remission of abdominal pain and complications. Moreover, appropriate therapeutic timing of ERCP and PTCD for obstructive SABP patients has not been established.

Thus, the efficacies of PTCD and ERCP in early treatment of obstructive SABP should be investigated. We retrospectively analyzed the clinical data for patients with obstructive SABP to evaluate the therapeutic effects of obstruction relieving by PTCD and ERCP as well as the therapeutic timing during interventions.

## 2. Methods

### 2.1. Patients and data collection

Patients from Beijing Tiantan Hospital who met the inclusion criteria (Table 1) were included in this study. The institutional research ethics committee of Beijing Tiantan Hospital, Capital Medical University approved this study. Clinical data for 62 patients with obstructive SABP were retrospectively analyzed at Beijing Tiantan Hospital, China, from July 2013 to July 2019. Based on treatments, 62 patients were assigned into three groups: Conservative (n=16), PTCD (n=22) and ERCP (n=24) groups. According to different treatment times, PTCD and ERCP groups were further assigned into early PTCD (⩽ 72 h, n=10) and delayed PTCD (>72 h, n=12) groups as well as early ERCP (⩽72 h, n=8) and delayed ERCP (>72 h, n=16) groups. Patients with ABP associated with cholangitis or bile duct obstructions^[23]^ and who had been subjected to ERCP were included in the ERCP group. Patients who could not tolerate endoscopy due to advanced age, persistent organ failure, multiple concomitant diseases, and clinical deterioration with signs or strong suspicion of infected necrotizing pancreatitis were included in the PTCD group. Patients who could not tolerate PTCD and ERCP/ENBD, as well as those who refused to accept any traumatic therapies were included in the Conservative group. In case the previously scheduled treatments failed, or in case of critical conditions, emergency interventions or surgical treatments were chosen with provision of a written informed consent^[24,25]^. The 62 SABP patients were finally enrolled in this study (Table 1; Fig. 1).

**Table 1:** Inclusion and exclusion criteria for SABP patients

| Criteria | Concomitant conditions or characteristics |
| --- | --- |
| Inclusion criteria | ● Acute pancreatitis<sup>[26]</sup><br>● Biliary cause of pancreatitis accompanied by POF (>48 h) (Organ failure was defined using the Modified Marshall scoring system)<sup>[27]</sup>; APACHE-II scores $\geq 8$; $\geq 3$ for Ranson; and $\geq 8$ for MCTSI<sup>[15]</sup><br>● Meet the determination criteria for biliary tract obstruction: **i.** Continuous increase in total bilirubin and direct bilirubin levels; **ii.** Imaging (abdominal ultrasound, CT, MRCP or EUS before ERCP and PTCD) suggesting biliary duct stones or total bile duct dilatation $\geq 1.0$ cm; **iii.** No obvious bile is introduced in gastrointestinal decompression. |
| Exclusion criteria | ● MABP (no evidence of organ dysfunction and without local and systemic complications; APACHE-II scores <8; <3 for Ranson and <4 for MCTSI)<br>● Combined with other serious diseases<br>● Pregnancy or puerperium<br>● Insufficient clinical data |

**Figure 1:**
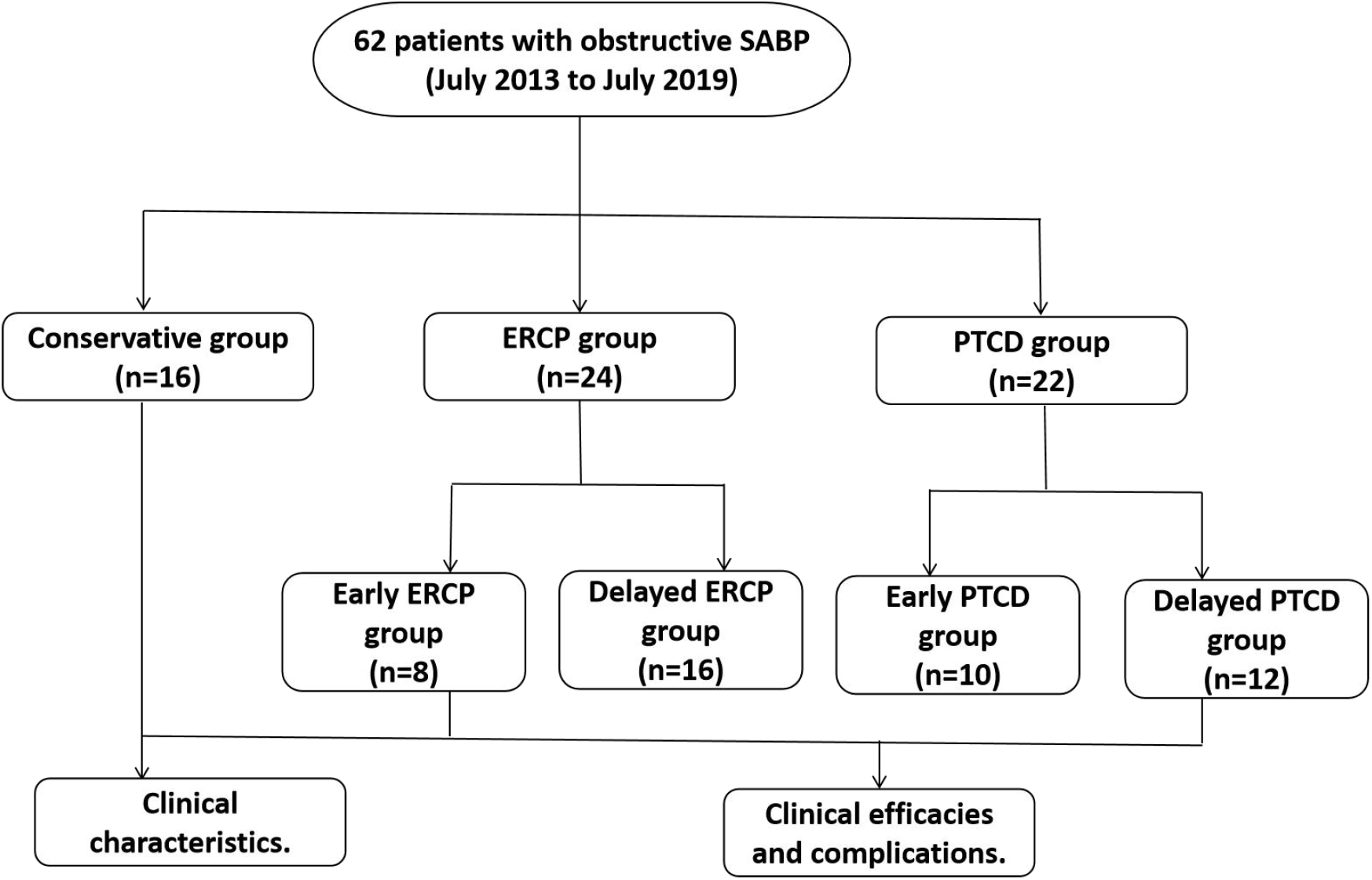
Flow diagram for this study.

### 2.2. Treatment procedures

#### 2.2.1. Conservative treatment

All patients in the conservative group were not subjected to ERCP or PTCD during admission, they were only conservatively treated, including: fasting for solids and liquids, liquid resuscitation, maintenance of hydroelectrolyte and acid-base balance, inhibition of pancreatic enzyme activities and pancreatic secretion, proton pump inhibitor for acid suppression, anti-inflammation if necessary and nutritional support.

#### 2.2.2. ERCP

In the ERCP group, patients accepted ERCP after admission either within 72 h or after 72 h. After completion of preoperative examination and other assessments, ERCPs were performed in a left-lateral position under sedation with alkaline 10 mg hydrochloride and 50 mg pethidine. Endocopy through the esophagus, stomach to descendant duodenum after which endoscopists adjusted the mirror body angle to expose the duodenal papilla. Then, retrograde bile duct intubation was performed, followed by slowly injecting iodinated contrast after successful operation to confirm whether the biliary tract had stones, stenosis or parapapillary diverticulum. If necessary, endoscopic sphincterectomy (ES), biliary lithotomy and endoscopic nasobiliary drainage (ENBD) were performed for management of choledocholithiasis.

#### 2.2.3. PTCD

Preoperative examinations for PTCD were similar to those of ERCP. Abdominal ultrasound was performed before operation to determine the bile duct or gallbladder puncture path and to locate the puncture point^[28]^. After intramuscular administration of 50 mg pethidine hydrochloride, routine disinfection and spreading towels, local infiltration anesthesia was performed. Under the guidance of ultrasound, all PTCDs were performed using an 18G puncture needle to target the bile tube or gallbladder. Bile was extracted after the needle had been removed, implying that the puncture had reached the target location. Then, a guide wire was inserted through the puncture needle. Using a subcutaneous catheter to dilate the puncture path, the 8F pigtail drainage tube was transplanted and the wire exited^[29,30]^. The drainage pipe was firmly secured to the skin and the sterile drainage bag externally attached.

### 2.3. Clinical characteristics

The following clinical indices of SABP groups were compared: recovery time for white blood cells, blood amylase and alanine transaminase (ALT), mean time of hospitalization and abdominal pain relief, probability of recovery and occurrence of complications. Complications that were compared included: acute accumulation of necrotic material, pancreatic pseudocyst, paralytic ileus, upper gastrointestinal hemorrhage, respiratory failure, systemic inflammatory response syndrome (SIRS), abdominal infection, sepsis, pancreatic encephalopathy and pancreatitis post-ERCP^[15]^.

### 2.4. Statistical analysis

Normally distributed continuous variables are expressed as mean ± standard deviation 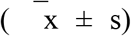. Measurement data between groups were compared using an independent sample *t*-test. Analysis of variance (ANOVA) was used to compare measurement data among groups. Enumerative data were expressed as rates or constituent ratios. Comparisons between groups were evaluated using *x*^*2*^-test and Fisher’s exact test. *P<*0.05 denoted significance. The SPSS software was used for statistical analyses.

## 3. Results

### 3.1. Data collection and clinical characteristics analysis

As shown in Table 2, clinical characteristics for the 62 patients with acute obstructive biliary pancreatitis were compared among conservative group, ERCP and PTCD groups. Most of the patients were female (55%) with an average age of 64 years. The levels of WBC, blood and urine amylase, ALT and TBiL were higher, specifically, ALT levels were more than three times the upper limit of normal. APACHE-II scores in the three groups were 16.06±4.99, 14.83±4.19, 15.32±4.58 (*p*>0.05), as well as Ranson score (4.63±0.81, 4.50±1.14, 4.41±1.26). Differences in complications such as previous attack of AP, cholelithiasis, heart disease, hypertension, diabetes, APACHE-II, Ranson and MCTSI scores among the three groups were not significant.

**Table 2:** Clinical data for obstructive SABP patients in the conservative, ERCP and PTCD groups

| Items | Conservative group (n=16) | ERCP (n=24) | PTCD (n=22) | F/$\chi^2$ 值 | p value |
| --- | --- | --- | --- | --- | --- |
| Age (years) | 62.81±18.42 | 65.25±16.49 | 64.18±17.15 | 0.096 | 0.908 |
| Gender [n (%)] |  |  |  |  |  |
| Male | 8 (50.00) | 11 (45.83) | 9 (40.91) | 0.316 | 0.854 |
| Female | 8 (50.00) | 13 (54.17) | 13 (59.09) | 0.316 | 0.854 |
| WBC (*10<sup>9</sup>/L) | 15.31±3.17 | 16.39±3.95 | 16.33±4.14 | 0.448 | 0.641 |
| Blood amylase (U/L) | 1241.69±422.23 | 1286.32±327.41 | 1209.80±278.67 | 0.296 | 0.745 |
| Urine amylase (U/L) | 3861.36±1283.74 | 3768.74±1402.64 | 3870.70±1283.55 | 0.165 | 0.849 |
| ALT (U/L) | 259.61±115.36 | 253.71±102.20 | 248.51±19.79 | 0.054 | 0.947 |
| TBIl (umol/L) | 41.66±12.72 | 43.86±13.31 | 42.96±15.28 | 0.120 | 0.887 |
| Complications [n (%)] |  |  |  |  |  |
| Previous attack of AP | 3 (18.75) | 4 (16.67) | 3 (13.64) |  | 0.913 |
| Cholelithiasis | 3 (18.75) | 9 (37.50) | 7 (31.82) |  | 0.505 |
| Heart disease | 5 (31.32) | 4 (16.67) | 4 (18.18) |  | 0.533 |
| Hypertension | 8 (50.50) | 9 (37.50) | 6 (27.27) | 2.053 | 0.358 |
| Diabetes | 2 (12.5) | 5 (20.83) | 4 (18.18) |  | 0.914 |
| APACHE-II score | 16.06±4.99 | 14.83±4.19 | 15.32±4.58 | 0.352 | 0.705 |
| Ranson score | 4.63±0.81 | 4.50±1.14 | 4.41±1.26 | 0.174 | 0.840 |
| MCTSI score | 7.19±0.83 | 6.92±0.78 | 6.91±0.75 | 0.731 | 0.486 |

### 3.2. Clinical efficacies and complications

The clinical efficacy, recovery rate and complications in the three groups as well as clinical indices in early and delayed groups were as shown in Tables 3, 4 and 5. The average length of stay for the conservative, ERCP and PTCD groups were 28.38±4.54, 16.71±3.99, 19.64±4.27, respectively. ERCP was most effective in shortening the duration of obstructive severe acute biliary pancreatitis (16.71 ± 3.99). Abdominal pain relief time, leukocyte remission time and recovery time of blood amylase, ALT and TBiL were shorter in ERCP and PTCD groups (*p*<0.05). Recovery rate of the conservative group was lowest (62.5%), when compared to ERCP (75.0%) and PTCD (86.36%) groups. Complications such as Pancreatic pseudocyst, acute accumulation of necrotic material and Sepsis et. al were lower in ERCP and PTCD groups, compared to the conservative group. Further, incidences of complications in the PTCD group significantly differed from those in the conservative group. Conservatively treated patients had high incidences of acute accumulation of necrotic material, pancreatic pseudocyst and sepsis.

**Table 3:** Clinical efficacies for obstructive SABP patients in the conservative, ERCP and PTCD groups

| Items (d) | Conservative group | ERCP | PTCD |
| --- | --- | --- | --- |
| Average length of stay | 28.38±4.54 | 16.71±3.99*<sup>#</sup> | 19.64±4.27* |
| Abdominal pain relief time | 11.93±2.26 | 8.15±4.24* | 8.25±2.94* |
| Leukocyte remission time | 14.62±3.91 | 8.94±3.70* | 9.27±2.64* |
| Blood amylase recovery time | 12.75±2.41 | 7.38±3.69* | 7.68±2.32* |
| ALT recovery time | 19.38±4.03 | 11.83±3.42* | 12.86±3.87* |
| TBiL recovery time | 17.55±3.86 | 10.92±2.901* | 12.61±3.781* |

\*ERCP and PTCD group compared to conservative group ( $p<0.05$ ).
<sup>#</sup>ERCP group compared to PTCD group ( $p<0.05$ ).

**Table 4:** Recovery rate and complications for obstructive SABP patients in the conservative, ERCP and PTCD groups

| Index | Conservative group (n=16) | ERCP (n=24) | PTCD (n=22) |
| --- | --- | --- | --- |
| Automatic discharge [n (%)] | 2 (12.50) | 2 (8.33) | 1 (4.55) |
| Recovery cases [n (%)] | 10 (62.50) | 18 (75.00) | 19 (86.36) |
| Complications cases [n (%)] | 10 (62.50) | 8 (33.33) | 6 (27.27) * |
| Acute accumulation of necrotic material (n) | 5 | 3 | 3 |
| Pancreatic pseudocyst (n) | 1 | 0 | 0 |
| Paralytic ileus (n) | 2 | 1 | 1 |
| Upper gastrointestinal hemorrhage (n) | 1 | 0 | 1 |
| Respiratory failure (n) | 3 | 3 | 2 |
| Systemic inflammatory response syndrome (n) | 8 | 8 | 6 |
| Abdominal infection (n) | 2 | 0 | 1 |
| Sepsis (n) | 1 | 0 | 0 |
| Pancreatic encephalopathy (n) | 1 | 0 | 0 |
| Cholangitis post-ERCP (n) | 0 | 1 | 0 |

\*PTCD group compared to the conservative group ( $P<0.05$ )

**Table 5:** Clinical efficacy indices and complications for obstructive SABP patient in early and delayed ERCP/PTCD groups

| Index | Early ERCP<br>(n=8) | Delayed ERCP<br>(n=16) | Early PTCD<br>(n=10) | Delayed PTCD<br>(n=12) |
| --- | --- | --- | --- | --- |
| Average length of stay (d) | 13.88±3.27<sup>* ^</sup> | 18.63±4.06<sup>+</sup> | 16.38±3.66<sup>#</sup> | 21.93±4.30 |
| Abdominal pain relief time (d) | 5.63±2.37 | 8.22±3.13 | 6.80±2.67<sup>#</sup> | 10.54±4.19 |
| Leukocyte remission time (d) | 6.31±0.92<sup>* ^</sup> | 9.88±2.406 | 8.50±2.07<sup>#</sup> | 11.14±4.09 |
| Blood amylase recovery time (d) | 5.50±1.31<sup>*</sup> | 8.89±1.91 | 6.13±1.73<sup>#</sup> | 10.14±3.98 |
| ALT recovery time (d) | 9.63±2.56 | 12.25±2.59<sup>+</sup> | 10.13±3.20<sup>#</sup> | 14.85±3.85 |
| TBiL recovery time (d) | 9.13±1.98<sup>* ^</sup> | 11.81±2.90 | 10.85±3.11<sup>#</sup> | 14.08±3.76 |
| Complication cases [n (%)] | 3 (37.50) | 5 (31.25) | 3 (30.00) | 3 (25.00) |

\*Early ERCP group vs delayed ERCP group ( $p<0.05$ )
<sup>#</sup>Early PTCD group vs delayed PTCD group ( $p<0.05$ )
<sup>^</sup>Early ERCP group vs early PTCD group ( $p<0.05$ )
<sup>+</sup>Delayed ERCP group vs delayed PTCD group ( $p<0.05$ )

The average length of stay (13.88±3.27), recovery time of leukocytes (6.31±0.92) and TBiL (9.13±1.98) in the early ERCP group were shorter than in both the delayed ERCP and early PTCD groups (*p*<0.05). The average length of stay (16.38±3.66), abdominal pain relief time (6.80±2.67), recovery time of leukocytes (8.50±2.07), blood amylase (6.13±1.73), TBiL (10.13±3.20) and ALT (10.85±3.11) in the early PTCD group were shorter, compared to the delayed PTCD group (*p*<0.05). The average length of stay (18.63±4.06) and ALT recovery time (12.25±2.59) in the delayed ERCP group were shorter than in the delayed PTCD group ((21.93±4.30) (14.85±3.85)) (*p*<0.05).

## 4. Discussion

Acute biliary pancreatitis is a common health care challenge in real-world clinical setting^[31]^. This condition is majorly caused by gallstones obstructing the biliopancreatic duct; however, persistence obstruction of the ampulla can theoretically aggravate pancreatic inflammation and may cause cascade activation of cytokines that mediates SIRS, pancreas and pancreatic tissue inflammation edema and necrosis^[32,33]^. Therefore, obstructive SABP is a clinical emergency with mortality rates of about 15%^[34]^, requiring prompt recognition and timely intervention to limit morbidity and prevent mortality or recurrence. Studies^[22,35]^ have shown that relieving biliary obstruction is an important measure for reducing SABP risk, to delay disease progression and improve prognosis.

Biliary obstruction can be eliminated by ERCP, PTCD and laparoscopic cholecystectomy^[36]^. Although there is no universally accepted treatment approach for SABP, the step-up approach using endoscopic drainage or percutaneous has been demonstrated to produce superior outcomes^[37]^. First, the 2019 Guidelines for management of acute biliary pancreatitis^[38]^ recommend ERCP for patients with any of the three high-risk criteria of cholangitis, radiological evidence of calculi, and total bilirubin >4 mg/dl combined with bile duct dilation. A meta-analysis^[22]^ involving 519 patients with pancreatitis and biliary obstruction found that routine use of ERCP reduced local complications. Second, PTCD is a minimally invasive surgical method with high patient cooperation. It is often used as an alternative method for surgical decompression in critically ill patients to relieve bile duct pressure. In this study, we assessed the therapeutic effects of conservative treatment, ERCP and PTCD in obstructive severe acute biliary pancreatitis and the choosing time of ERCP and PTCD. We established that clinical efficacies of ERCP and PTCD for obstructive SABP were comparable and that both approaches are beneficial for recovery of laboratory indicators; patients subjected to ERCP or PTCD had shorter remission times of abdominal pain, hospital stay, lower incidences and mortality of sepsis. Complications due to ERCP and PTCD were fewer than those of conservative treatment. Our findings are consistent with those of previous studies^[21,22,34]^.

For all gallstone pancreatitis patients with suspected ascending cholangitis, prompt (within 48 h of presentation) ERCP is recommended. Given that there is no consensus on guidelines for obstruction relief of SABP and that emergency ERCP may not always be performed in clinical practice, we evaluated the therapeutic effects of early (⩽ 72 h) and delayed (>72 h) ERCP in ABP patients with cholangitis. In this study, clinical efficacies of early ERCP and PTCD were better than those of delayed ERCP and PTCD. Early endoscopic and interventional therapy is recommended for relieving obstruction of patients with obstructive severe acute biliary pancreatitis within 72 h after admission. Our findings provide an important indication for future prospective studies.

SABP should be managed by intensive monitoring and systemic support. Conservative treatment is aimed at achieving supportive therapy, resuscitation and to treat specific complications that may occur^[39,40]^. Conservative therapy is imperative and fundamental for SABP patients. In this study, the clinical efficacies and treatment satisfaction of patients subjected to conservative treatment alone were poor. Since the treatment was not feasible or satisfactory, three patients in the conservative group were eventually treated with ERCP. Due to critical conditions of obstructive SABP, clinicians may be concerned about the risk of conservative medical management. In conclusion, conservative therapy is the basis of obstructive SABP, but timely intervention is reasonable within 72 h for laboratory indices, hospitalization stays, recovery of liver function and remission of abdominal pain and complications. If early ERCP is not safe and feasible, PTCD should be performed and has certain clinical effect.

This study is associated with some limitations. First, surgical risks of patients were not assessed, which could have led to deviations in the success rates of surgeries, subsequent cure rates and postoperative complications among other aspects. Second, the sample size was limited to a single center retrospective study, which may have resulted in the absence of significant differences in recovery rate. Given these limitations, a stricter standard, follow-up visits, high-quality and large sample tests are required to verify the reliability of our conclusions. Further researches are needed and other researchers can use this research to obtain strategies for treatment and management of severe acute biliary pancreatitis.

## Conclusions

Removal of obstruction of SABP is a focus of clinical attention. Decision on the timing of procedural interventions is also crucial for improving morbidity or mortality outcomes. In summary, ERCP and PTCD are effective for relieving biliary obstruction of severe acute biliary pancreatitis. Early ERCP or PTCD (within 72 h) for obstructive SABP are more beneficial.

## Data Availability

All data produced in the present study are available upon reasonable request to the authors

## Abbreviations

PTCD: Percutaneous transhepatic cholangial drainage
ERCP: Endoscopic retrograde cholangiopancreatography
ABP: Acute biliary pancreatitis
SABP: Severe acute biliary pancreatitis
MABP: Mild acute biliary pancreatitis
SIRS: Systemic inflammation response syndrome
MODS: Multiple organ dysfunction syndrome
POF: Persistent organ failure

